# Machine Learning Classification of Mild Cognitive Impairment using Advanced Multi-Shell Diffusion MRI and CSF Biomarkers

**DOI:** 10.1101/2025.02.27.25322792

**Authors:** Alexander Y. Guo, John P. Laporte, Kavita Singh, Jonghyun Bae, Keagan Bergeron, Angelique de Rouen, Noam Y. Fox, Nathan Zhang, Isabel Carino-Bazan, Mary E. Faulkner, Dan Benjamini, Zhaoyuan Gong, Mustapha Bouhrara, the Alzheimer’s Disease Neuroimaging Initiative

## Abstract

**INTRODUCTION:** Machine learning applied to neuroimaging can help with medical diagnosis and early detection by identifying biomarkers of subtle changes in brain structure and function. The effectiveness of advanced diffusion MRI (dMRI) imaging methods for pre-dementia classification remains largely unexplored, particularly when combined with CSF biomarkers.

**METHODS:** We implemented XGBoost machine learning models to evaluate the classification potential of dMRI parameters (derived using NODDI, C-NODDI, MAP, or SMI), CSF biomarkers of Alzheimer’s pathology (*Tau*, *pTau*, *Aβ42*, *Aβ40*), and pairwise dMRI + CSF combinations in distinguishing cognitive normality from mild cognitive impairment.

**RESULTS:** MAP*-RTAP* (AUC=0.78) and *pTau/Aβ42* (AUC=0.76) were the best performing individual biomarkers. Combining C-NODDI*-C-NDI* and *Aβ42/Aβ40* achieved the highest performance (AUC=0.84) and accuracy (0.84), while other combinations optimized either sensitivity (0.93) or specificity (0.88).

**DISCUSSION:** dMRI biomarkers demonstrate comparable performance to CSF biomarkers, with notable improvements achieved when combined. This study highlights dMRI’s potential for enhancing early AD detection.

## 1. INTRODUCTION

A major goal in Alzheimer’s disease (AD) research is to identify highly sensitive and specific biomarkers for diagnosis, especially in its early stages. Growing attention has focused on mild cognitive impairment (MCI), a prodromal phase of AD where patients exhibit memory declines resembling AD [1, 2]. Notably, 12-20% individuals with MCI progress to dementia annually [3, 4]. Therefore, early and accurate MCI diagnosis is essential for timely interventions and treatment outcomes.

AD and MCI have historically been regarded as a gray matter (GM) disease, with cortical atrophy and ventricular enlargement on the macroscopic scale, and development of amyloid-β plaques and neurofibrillary tau tangles on the microscopic scale [5, 6]. As a result, CSF biomarkers are of prominent interest in the early detection and monitoring of AD and MCI. Levels of these proteins help identify AD-related changes before symptoms appear, providing a more objective diagnosis than cognitive tests alone. These biomarkers also assist in tracking disease progression and evaluating treatment effectiveness, enabling personalized care and better outcomes [7–9]. However, while their growing importance in both research and clinical settings supports earlier intervention attempts and advances our understanding of neurodegenerative diseases, the limitations of CSF biomarkers, such as their invasiveness, variability due to factors like age and comorbidities, and the need for additional clinical and imaging data for a complete diagnosis, must be carefully considered [10]. Moreover, in the recent years, it has been suggested that alterations in cerebral white matter (WM), including demyelination, axonal loss, and synaptic degeneration, may precede amyloid-β and tau protein deposition, while demonstrating much stronger relationships to dementias’ severity and cognitive deficits [11–13].

Concurrently, the last three decades have seen a myriad of advanced diffusion MRI (dMRI) methods to characterize cerebral WM microstructure with unprecedented sensitivity [14, 15]. Notably, diffusion tensor imaging (DTI) [16, 17], which describes the distribution of diffusion displacements using a statistical Gaussian model, was used in extensive works to demonstrate age-differentiated trajectories of cerebral WM tissues during brain maturation and degeneration [15, 16, 18]. However, molecular water diffusion in brain follows a non-Gaussian distribution due to the restriction of cell membranes, organelles, and water compartments. Therefore, despite the promising sensitivity of DTI, it has inherent limiting assumption hampers utility and specific interpretation [19, 20]. To improve specificity, biophysical diffusion approaches have been introduced, namely the Neurite Orientation Distribution and Density Imaging (NODDI) [21, 22], Constrained NODDI (C-NODDI) [23], and Standard Model Imaging (SMI) [24–26]. These models employ a multi-compartment framework to estimate biophysical parameters, such as neurite density and axonal fraction, offering greater specificity to the underlying brain microstructure and composition at the cost of far greater estimation complexity. To achieve a balance between the highly sensitive, but simplistic, DTI modeling, and the more specific, but mathematically intricate NODDI/C-NODDI/SMI models, the mean apparent propagator (MAP) model was also introduced [27]. The MAP model takes a unique approach by avoiding Gaussian assumptions about water behavior in tissues and instead employs a fully probabilistic framework to evaluate molecular dispersion, enabling an assessment of water movements. MAP metrics include the return-to-origin probability (RTOP), a measure of the likelihood of water molecules undergoing zero net displacements due to restricting barriers, as well as the return-to-axis probability (RTAP) and the return-to-plane probability (RTPP), measures of the presence of restrictive barriers in the radial and axial orientations, respectively. In addition, the non-Gaussianity index (NG) metric reflects the extent of deviation from a homogeneous/Gaussian diffusivity, and the propagator anisotropy (PA) provides an indicator of the anisotropy and dispersion of the fibers. Consequently, MAP statistically captures the microstructural complexity of cerebral tissues encompassing a wide range of underlying structural and architectural characteristics, at the cost of lacking biologically relevant specificity of the cerebral tissue being investigated. Both biophysical models like NODDI and SMI, as well as statistical ones like MAP, are increasingly used to study brain aging and neurodegeneration [28–32], though these studies have been sparse and are often conducted on cohorts with limited sample sizes.

The main goals of this study are to assess the classification performance of NODDI, C-NODDI, MAP, and SMI parameters in WM, and to compare the results to performance of CSF biomarkers of AD pathology, namely, *Tau*, *pTau*, *Aβ_40_*, and *Aβ_42_*. Additionally, we assessed the classification performance when using both dMRI and CSF biomarkers simultaneously. Our machine learning framework of choice was eXtreme Gradient Boosting (XGBoost), an algorithm known for its strong performance in neuroimaging classification and an ability to highlight important features [33–36]. XGBoost builds multiple decision trees in an iterative process, improving accuracy by learning from mistakes made in previous steps. It also includes techniques to prevent overfitting, through limiting complexity and selecting only the most useful information from the data, allowing it to work well even with complex and limited datasets. We conducted our analysis using CSF data and multi-shell dMRI images collected during the third phase of the Alzheimer’s Disease Neuroimaging Initiative (ADNI 3).

## **2.** METHODS

### ***2.1.*** Participants

Participants were drawn from the ADNI database (https://adni.loni.usc.edu/). The ADNI initiative was started in 2003 under Michael W. Weiner, MD, and was a private-public partnership funded by private companies as well as the NIH and NIA for the purpose of developing clinical, imaging, genetic, and biochemical biomarkers for early detection of Alzheimer’s Disease (AD). Inclusion criteria were participants who had multi-shell diffusion scans on Siemens scanners and CSF biomarker data available in ADNI 3 as of January 3^rd^, 2024. Baseline dMRI scans were obtained across all available participants (*n* =189), of which 114 were cognitively normal (CN) while 75 had MCI (Table 1). A subset of these participants also had fluid biomarkers (Table 1). All participants signed consent forms, and study design was approved by the IRB of data-collection institutions. For more information, see www.adni-info.org.

**Table 1:**
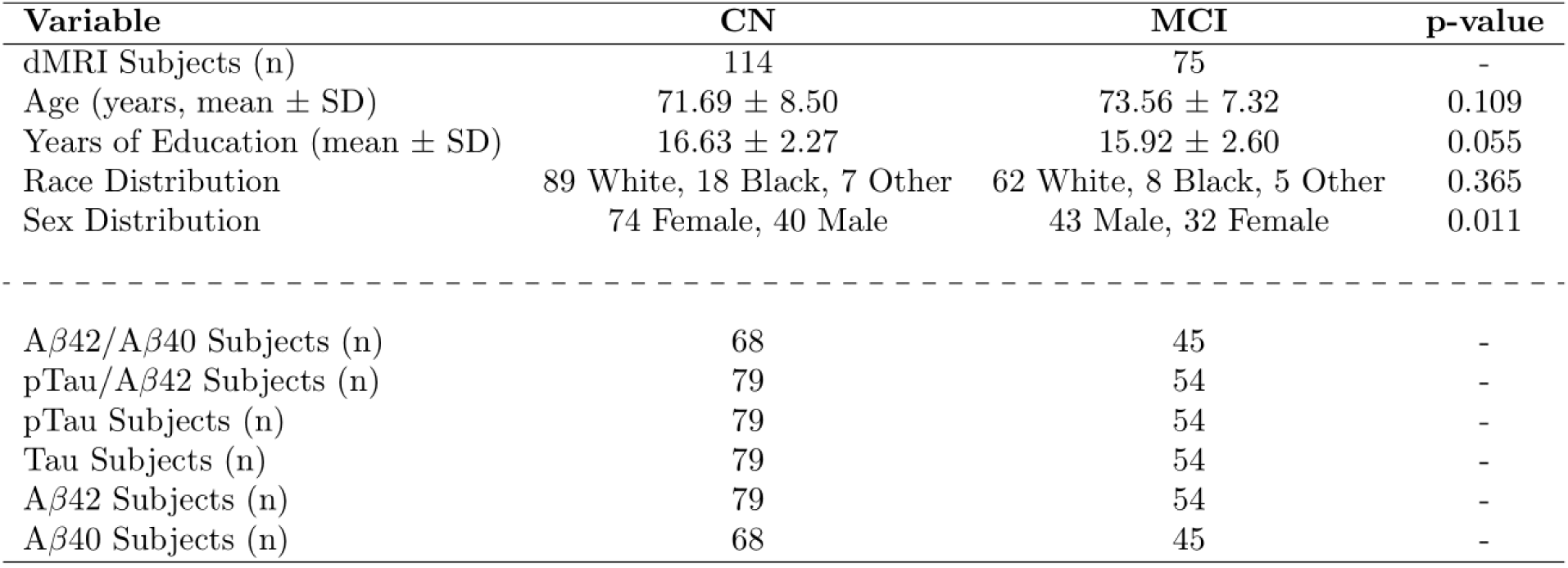
Demographic and biomarker sample size of the study cohort.

| Variable | CN | MCI | p-value |
| --- | --- | --- | --- |
| dMRI Subjects (n) | 114 | 75 | - |
| Age (years, mean ± SD) | 71.69 ± 8.50 | 73.56 ± 7.32 | 0.109 |
| Years of Education (mean ± SD) | 16.63 ± 2.27 | 15.92 ± 2.60 | 0.055 |
| Race Distribution | 89 White, 18 Black, 7 Other | 62 White, 8 Black, 5 Other | 0.365 |
| Sex Distribution | 74 Female, 40 Male | 43 Male, 32 Female | 0.011 |
| ----- |  |  |  |
| Aβ42/Aβ40 Subjects (n) | 68 | 45 | - |
| pTau/Aβ42 Subjects (n) | 79 | 54 | - |
| pTau Subjects (n) | 79 | 54 | - |
| Tau Subjects (n) | 79 | 54 | - |
| Aβ42 Subjects (n) | 79 | 54 | - |
| Aβ40 Subjects (n) | 68 | 45 | - |

### ***2.2.*** dMRI Imaging Protocol

Each participant underwent whole brain dMRI scans using a 3 T Siemens Prisma VE11C scanner. Multi-shell diffusion MRI data were collected with a repetition time of 3400 ms and an echo time of 71 ms. Each dMRI scan included 127 separate images: 13 with *b* = 0 s/mm², 6 with *b* = 500 s/mm², 48 with *b* = 1000 s/mm², and 60 with *b* = 2000 s/mm². 15 raw dMRI scans were eventually discarded due to incomplete fields of view or severe motion artifacts.

### ***2.3.*** Image Pre-Processing

dMRI scans were pre-processed and extracted for region-of-interest (ROI) values through an eight-step pipeline before classification (Figure 1). First, the raw imaging dataset was processed in Python to convert individual DICOM images into 4-D NIfTI files and standardized in format. Images were subsequently denoised using the MP-PCA MATLAB toolbox [37]. Brain extraction and linear-registration of each diffusion-weighted image to the non-diffusion weighted b_0_ image was performed using the FMRIB Software Library (FSL) [38]. Eddy current-induced distortions and participant movements were corrected for using FSL, and dMRI imaging biomarkers were then derived using NODDI, C-NODDI, MAP, and SMI models. Lastly, derived parameter maps were registered to the JHU-ICBM-T_2_-2mm template using linear and nonlinear FSL FLIRT and FNIRT functions before ROI analysis was conducted using the JHU DTI atlases. Non-white matter ROI voxels were removed based on a threshold of a WM probability less than 0.95, and mean ROI values were then calculated for the frontal, temporal, parietal, and occipital lobes, as well as for the whole brain and cerebellum.

**Figure 1:**
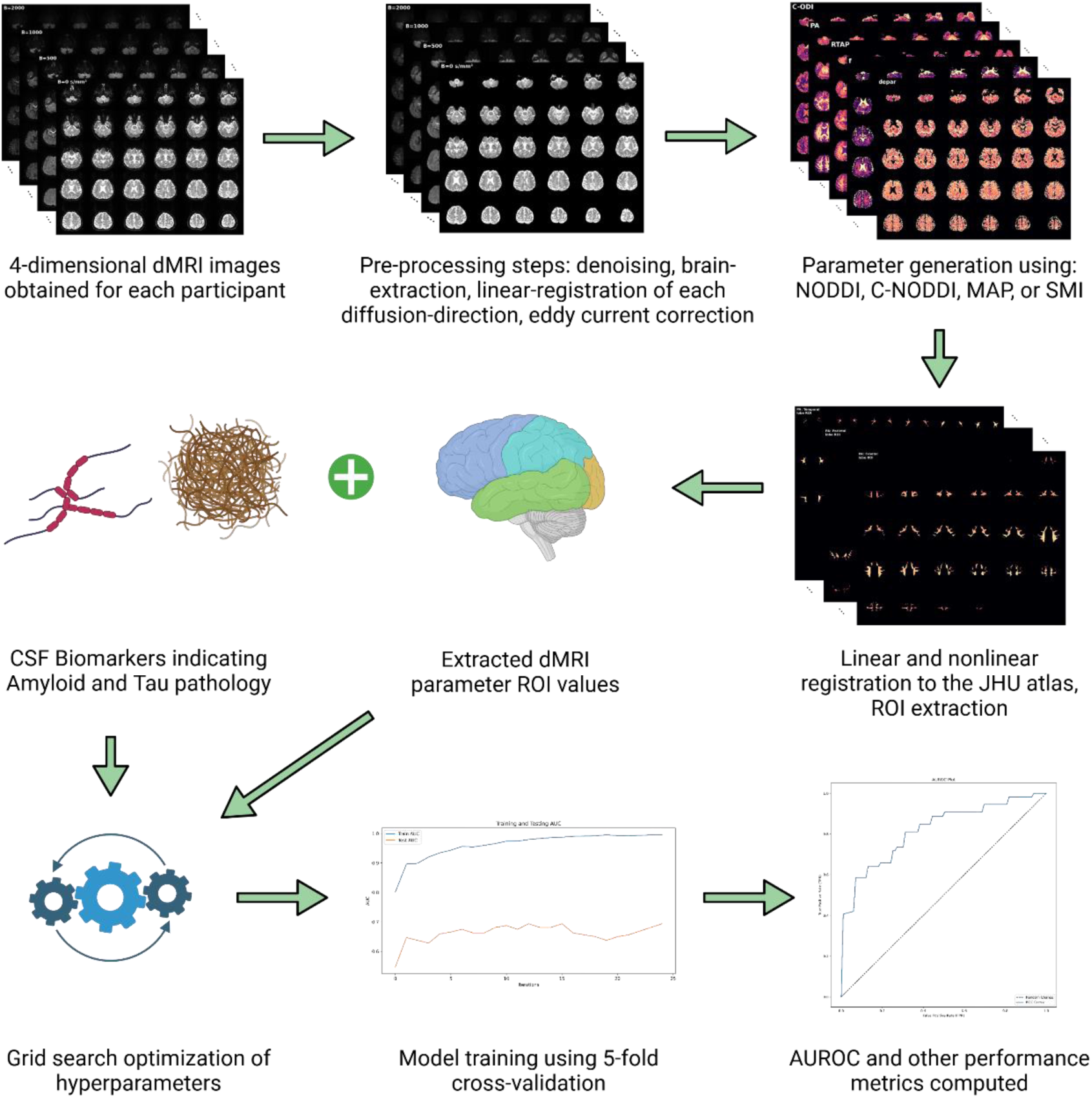
Image preprocessing and classification pipeline.

### ***2.4.*** dMRI Biomarkers

In total, 14 dMRI biomarkers were estimated for each participant. Two parameters were derived using NODDI or C-NODDI, namely, NODDI-*ODI* (orientation dispersion index), NODDI-*NDI* (neurite density index), C-NODDI-*C-ODI* (constrained-ODI), and C-NODDI-*C-NDI* (constrained-NDI). Five parameters were obtained using the MAP model, namely, MAP-*NG* (non-Gaussianity), MAP-*PA* (propagator anisotropy), MAP-*RTOP* (return to origin probability), MAP-*RTPP* (return to plane probability), and MAP-*RTAP* (return to axis probability). Five were derived using the SMI model, namely, SMI-*ƒ* (axon fraction), SMI-*D_a_* (intracellular axial diffusion), SMI-*D_e,_*_‖_ (extracellular parallel diffusion), SMI-*D_e,_*_〦 (extracellular perpendicular diffusion), and SMI-_*p_2_* (2^nd^ order of the orientation distribution function).

### ***2.5.*** CSF Biomarkers

Lumbar CSF samples were collected and stored in −80°C pending analyses at the ADNI Biomarker Core in the University of Pennsylvania School of Medicine. The 4 established CSF biomarkers of amyloid-*β*40 *(Aβ40)*, amyloid-*β*42 *(Aβ42)*, Tau protein (*Tau)*, and phosphorylated-Tau *(pTau)* were obtained from a subset of ADNI participants who had dMRI scans (*n* = 133). *Aβ42/Aβ40* and *pTau/Aβ42* ratios were subsequently computed for each participant. Further collection details can be found through ADNI Documentation (https://adni.loni.usc.edu/help-faqs/adni-documentation/).

### ***2.6.*** Machine Learning Classification

Diagnostic classification was performed in Python using XGBoost [36] (Figure 1). Models classifying participants into CN versus MCI were optimized for each individual MRI parameter, each individual CSF biomarker, and each MRI + CSF biomarker combination. Feature selection of the MRI parameters was first performed using XGBoost feature importance explainability plots with minimally contributing features (race, acquisition site ID, occipital lobe, cerebellum) removed. The whole brain ROI was also removed due to high correlation with the other remaining ROIs. The final subset of demographic information used were age, sex, and years of education (YoE), while the frontal, temporal, and parietal lobes became the final dMRI ROIs. Cross-validated grid search was then performed for hyperparameter tuning of each model [39]. 5-fold cross-validation [40] with an 80/20 split was used for model training, with SMOTE [41] oversampling performed to balance class labels. Test performance was compared using the area under the curve (AUC) of the receiver operating characteristic (ROC) [42], and other performance metrics (accuracy, precision, recall, F1-score, sensitivity, specificity) were computed over the test set of each fold, then averaged across the 5 folds. An optimal threshold for each classifier was calculated using Youden’s J statistic [43] when computing performance metrics. To interpret the model’s predictions, SHAP (SHapley Additive exPlanations) [44] analyses was conducted to visualize feature importance and contributions to model decisions.

## 3. RESULTS

### ***3.1.*** Group Comparisons of the dMRI-derived and CSF Biomarkers

Violin plots were used to visualize the distribution of the CSF and dMRI-derived parameters across diagnostic groups (CN versus MCI) in the frontal, parietal, and temporal WM lobes, with wider sections representing a higher density of data points. Independent two-sample *t*-tests were conducted to assess differences between the two groups for each dMRI biomarker, with significance thresholds set at *p* < 0.05. Figure 2a shows the six dMRI-derived parameters that exhibited significant differences between CN and MCI across all three of the frontal, parietal, and temporal lobe WM, indicating their potential strength in downstream classification tasks. These parameters included C-NODDI-*C-NDI*, MAP-*PA*, MAP-*RTPP*, MAP-*RTOP*, MAP-*RTAP*, and SMI-*D_e,_*_〦_. Other dMRI parameters had fewer than 3 significant ROIs.

**Figure 2a:**
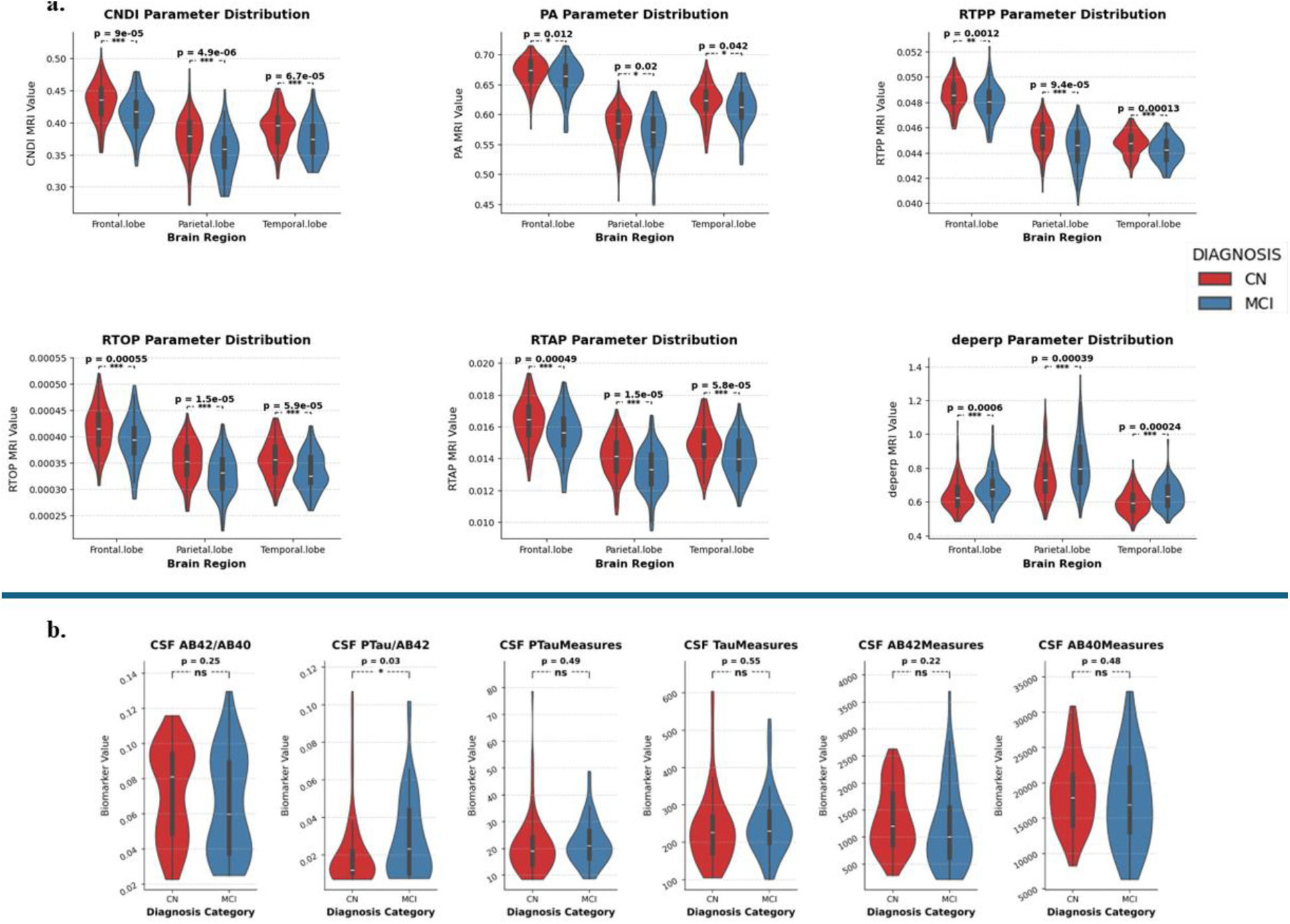
Violin plots showing the dMRI parameter distribution values for three selected ROIs. Only dMRI parameters with at least three significant ROIs are displayed. Figure 2b: Violin plots showing CSF biomarker distributions. Significance levels are indicated above each plot, with “ns” denoting non-significance.

The distribution of CSF biomarker values for CN and MCI groups are shown in Figure 2b. Among the biomarkers, only *pTau*/*Aβ42* showed a statistically significant difference between CN and MCI groups (*p* = 0.03), indicating potential discriminative power for MCI classification. In contrast, the other biomarkers, including *Aβ42*/*Aβ40*, *pTau*, *Tau*, *Aβ42*, and *Aβ40* did not show statistically significant differences between the groups (*p* > 0.05).

### ***3.2.*** Classification Performance of dMRI and CSF Biomarkers

Figure 3a displays the ROC for the top six dMRI or CSF biomarkers in distinguishing between CN and MCI participants. Of the dMRI parameters, MAP-*RTAP* demonstrated the highest classification performance (AUC = 0.78), followed by SMI-*D_e,_*_〦 (AUC = 0.76), MAP-*RTPP*, MAP-*PA*, and MAP-*RTOP* (AUC = 0.74), and then C-NODDI-*C-NDI* (AUC = 0.73). Among CSF_ biomarkers, the *pTau*/*Aβ42* ratio performed best (AUC = 0.76), with the other CSF biomarkers following relatively closely (AUCs ≤ 0.73).

**Figure 3a:**
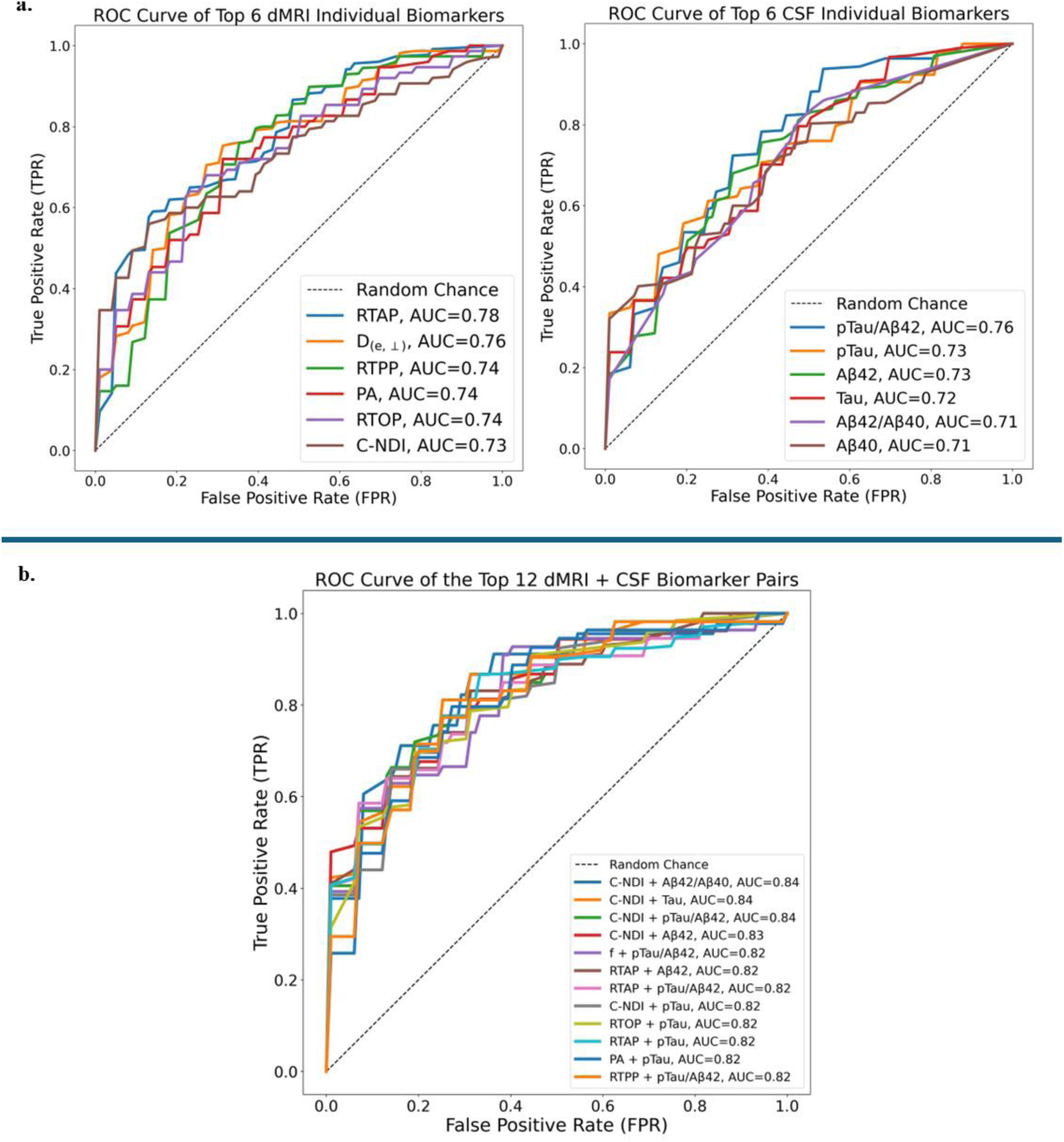
ROC displaying the AUC performance of each classifier using patient demographics (age, sex, YoE) in combination with dMRI parameters (left) or CSF biomarker values (right). Only the top 6 performing classifiers are shown in each plot. Figure 3b: ROC displaying the top 12 classifiers utilizing patient demographics (age, sex, YoE) with dMRI + CSF biomarker combinations. All dMRI parameters utilized ROI information from the frontal, parietal, and temporal lobes.

Figure 3b presents the ROC for the top 12 dMRI-CSF combined biomarker pairs. Combining dMRI and CSF biomarkers led to marked improvements in AUC across the board. The highest performances were obtained with the C-NODDI-*C-NDI* parameter when combined with either *Aβ42*/*Aβ40*, *Tau*, *pTau*, or *Aβ42* (AUCs = 0.84, 0.84, 0.84, 0.83). This was followed by the SMI-*f* parameter when combined with *pTau*/*Aβ42* (AUC = 0.82). The other classifiers were MAP-*RTAP*, MAP-*RTOP,* MAP*-PA and* MAP*-RTPP* parameters when combined with various CSF biomarkers (AUC = 0.82).

### ***3.3.*** Performance Metrics

The classification performance of individual dMRI parameters and CSF biomarkers are summarized in Table 2a. Among the biomarkers with the highest AUCs, C-NODDI-*C-NDI* demonstrated the best overall performance, achieving an accuracy of 0.78 and an F1-score of 0.76, alongside a low sensitivity (0.53) but an outstanding specificity (0.94). In contrast, MAP-*RTAP* showed similar accuracy and F1-score while displaying the opposite sensitivity-specificity trade-off, with higher sensitivity (0.84) and lower specificity (0.68). Among CSF biomarkers, *pTau*/*Aβ42* emerged as the top classifier, performing comparably to MAP-*RTAP* with a sensitivity of 0.92 and specificity of 0.61.

**Table 2a:**
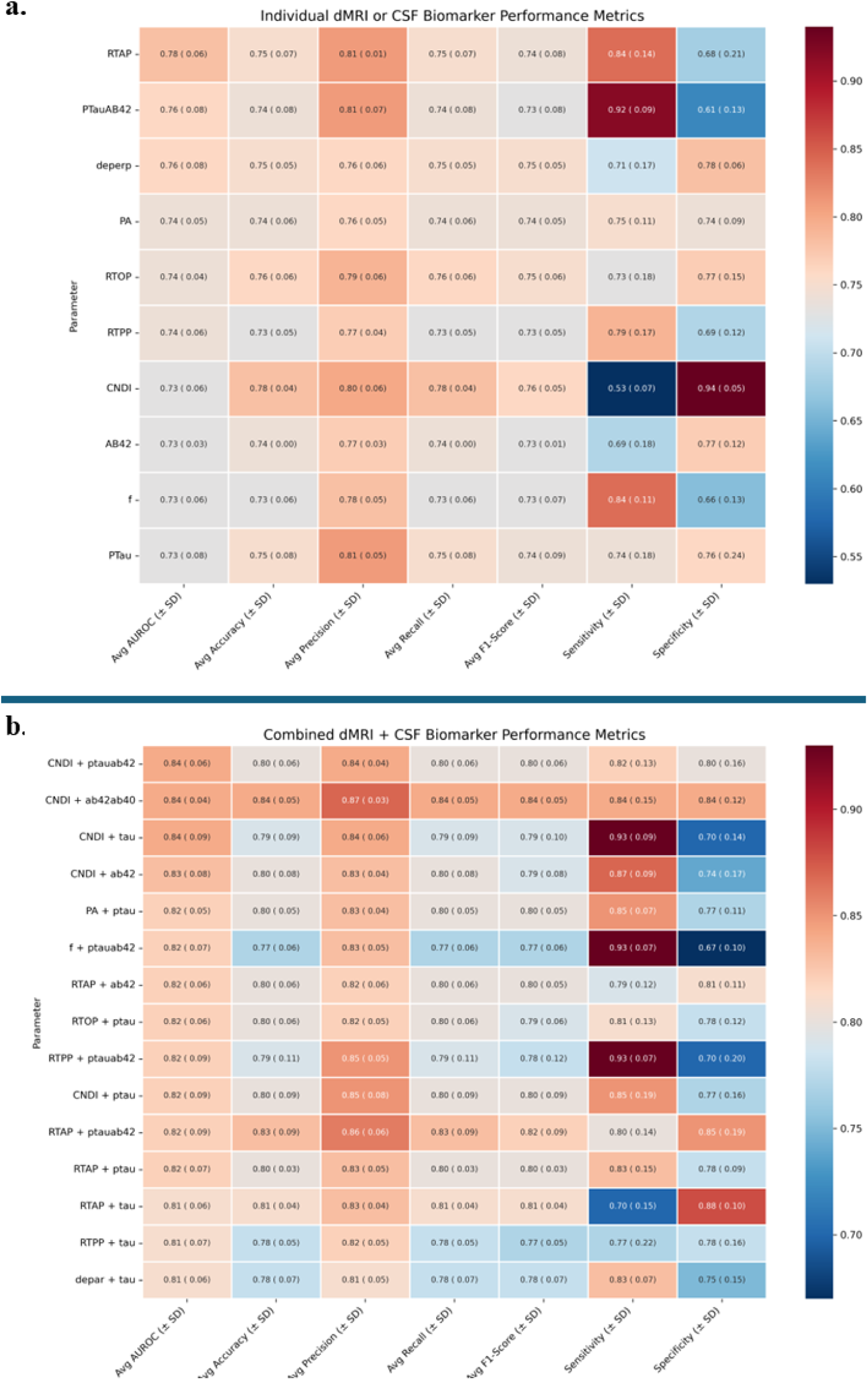
Test set performance metrics of each of the individual biomarker models, averaged across cross-validation folds. Parameters ordered top to bottom in decreasing AUROC value, with the top 10 models shown. Table 2b: Performance metrics of the combined dMRI + CSF biomarker models. Only the top 15 models are shown.

The combination of dMRI and CSF biomarkers led to noticeable improvements compared to individual biomarkers alone (Table 2b). The highest-performing combinations, based on AUC, all demonstrated strong accuracy (≥ 0.79) and F1-score (≥ 0.79). C-NODDI-*C-NDI + Aβ42/Aβ40* emerged as the top-performing model, achieving an accuracy and F1-score of 0.84, along with a well-balanced sensitivity and specificity of 0.84 each. Other notable combinations included C-NODDI-*C-NDI* + *Tau* which achieved high sensitivity (0.93) but moderate specificity (0.70), and MAP-*RTAP* + *Tau* which exhibited the opposite balance, with moderate sensitivity of 0.70 and a high specificity of 0.88.

### ***3.4.*** SHAP Analysis: Impact of Features on Model Predictions

To gain insights into how the XGBoost models made predictions, we conducted SHAP analysis, which quantifies feature contributions by computing Shapley values originating from cooperative game theory. Specifically, for a given participant, SHAP evaluates the difference in model output with and without the feature, averaging over all possible subsets of the feature set to ensure a fair attribution of importance while maintaining additivity, consistency, and local accuracy. Figure 4 displays input features ranked by their importance, determined by the average improvement in model accuracy when a feature is used in a split, along the y-axis. Additionally, each point within a row represents an individual participant, with red indicating higher feature values and blue indicating lower values. The x-axis represents the magnitude and direction of each feature’s influence on the model’s output predictions. Negative SHAP values on the x-axis are associated with a higher likelihood of CN classification, while positive SHAP values indicate a greater likelihood of MCI classification.

**Figure 4:**
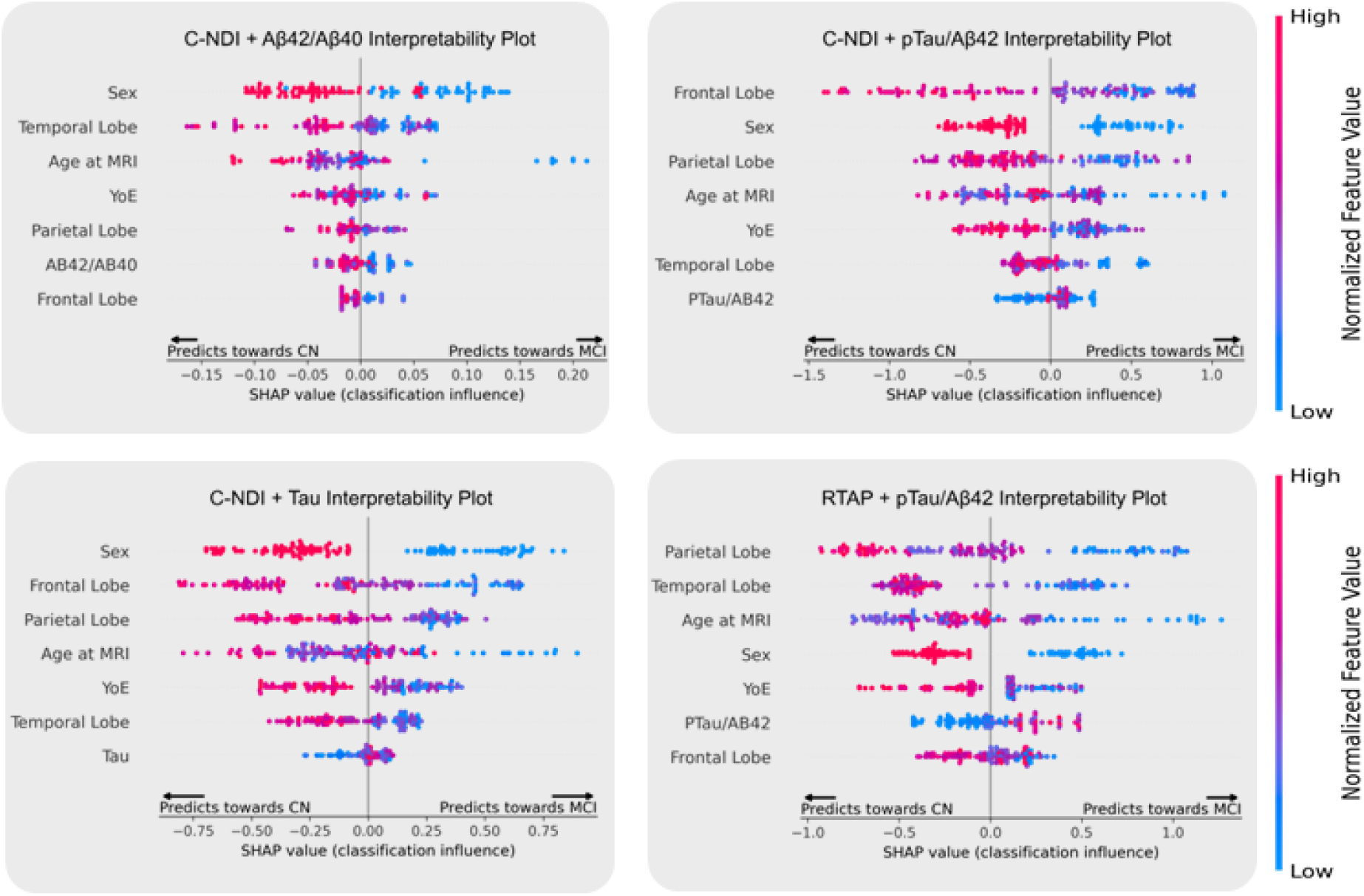
Feature importance and SHAP values for four representative combined models. Sex is binary-encoded, with males assigned a value of 0 and females a value of 1.

**Figure 5:**
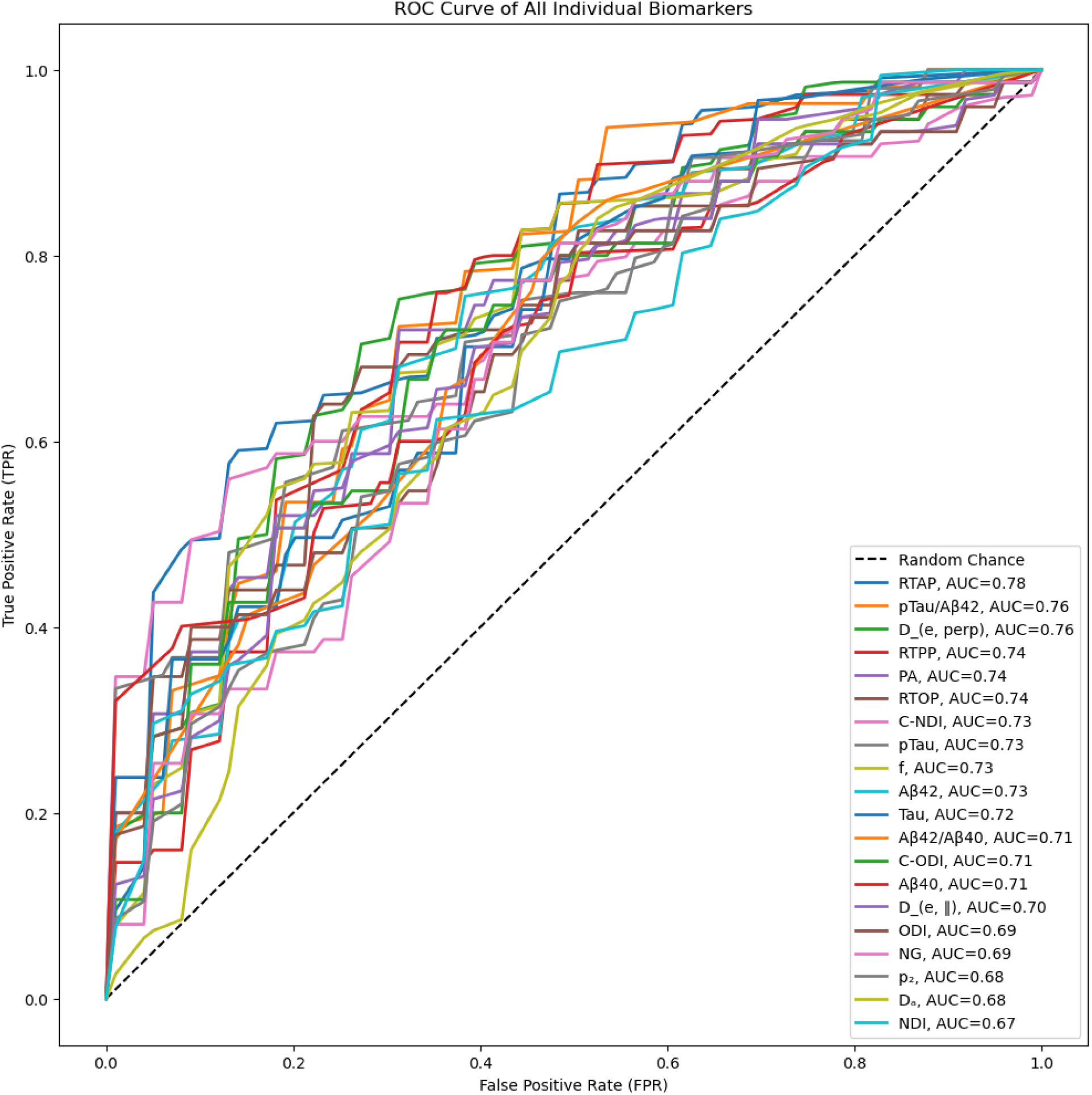
ROC curves displaying AUC performance of all individual classifiers using patient demographics (age, sex, YoE) in combination with dMRI parameters or CSF biomarker values.

In each representative model, at least two of the three WM lobe ROIs were among the most influential features, appearing near the top of the y-axis. Lower dMRI ROI values (blue points) were generally associated with a higher likelihood of MCI classification, as indicated by positive SHAP values. The demographic feature of sex also had fairly high importance and showed a clear pattern, with male sex (blue points) strongly linked to MCI. Meanwhile, age had moderate importance, with a complex relationship that showed both high and low feature values contributing to CN classification, while a few low valued points distinctly associated with MCI. Lastly, YoE and CSF biomarkers were generally of lower importance but showed clear stratification, where low YoE and *Aβ42*/*Aβ40* ratios, as well as high *pTau/Aβ42* ratios and *Tau* values, were linked to MCI diagnosis.

## 4. DISCUSSION

In this study, we assessed the performance of dMRI or CSF biomarkers, as well as their pairwise combinations, in distinguishing between CN and MCI diagnoses. Our findings reveal that several dMRI-derived parameters, particularly those from the statistical MAP model, exhibit significant differences between clinical groups, and yield strong ML classification performance. While CSF biomarkers exhibited slightly lower performance, integrating dMRI and CSF biomarkers together significantly improved all metrics, with notable benefit to the C-NODDI-*C-NDI* parameter. These results highlight the complementary nature of dMRI and CSF modalities in identifying MCI-related brain differences.

### ***4.1.*** Individual Biomarker Comparisons

The dMRI parameter distributions in WM readily show significant (p < 0.05) differences between CN and MCI individuals, highlighting the presence of early WM changes associated with MCI. This aligns with previous studies indicating that changes to brain regions such as the medial temporal lobe and posterior cingulate/precuneus networks are crucial in early AD and may contribute to cognitive decline [45, 46]. The dMRI parameters with significant group differences coincided with the dMRI parameters exhibiting the highest classification success. Of these, MAP-*RTAP*, which is sensitive to restrictive barriers perpendicular to the principal direction of diffusion, showed the highest AUC performance, with MCI showing significantly reduced MAP-*RTAP* values as compared to CN (Figure 2). Previous studies have reported that MAP-*RTAP* is inversely related to the apparent diameter of the axon [27, 47, 48] and therefore, the reduction in MAP-*RTAP* may be due to an increase in axonal diameter as a result of probable demyelination or axonal swelling. This was further supported by the high performances by SMI-*D_e,_*_〦 and related MAP-derived metrics, namely, MAP-*RTPP*, MAP-*PA*, and MAP-*RTOP*, which all further indicate changes to myelin content through their respective sensitivities to alterations in diffusion properties. Collectively, these results indicate the importance of using WM microstructural changes and diffusion restriction in identifying early changes associated with MCI._

Furthermore, our study provides the first concrete evidence that the MAP parameters in WM, when used without CSF information, are overwhelmingly better suited for classification of MCI compared to those derived using multi-component biophysical models such as NODDI and SMI. MAP is fitted to overall diffusion signals (from both intra- and extra-cellular compartments) and imposes few assumptions, potentially offering greater sensitivity than multi-compartment models. The prevalence of MAP parameters among the best individual classifiers also agrees with a recent study that found them to be more sensitive to AD pathology and neurodegeneration than NODDI and DTI [32]. On the other hand, SMI parameters, besides *D_e,_*_〦_, obtained relatively lower classification performance. This may be due to SMI’s approach of avoiding incorporating fixed parameters to reduce fitting degeneracy, and its attempt to determine more than five parameters at once, including axonal water fraction, a surrogate of axonal density, as well as intra- and extra-axonal diffusivities, proxies of myelin status and axonal integrity. Such an approach involves a difficult high-dimensional inverse problem to solve, and therefore, despite its potential added specificity, derived parameters are dramatically dependent on very complex underlying assumptions and increasingly sensitive to the impact of experimental noise, potentially resulting in the low performance seen in this study. A thorough follow-up investigation of the performance of SMI derived parameters for MCI classification is still required, ideally with multi-shell dMRI data acquired with more or higher b-values than those incorporated in ADNI 3. Like SMI, NODDI is a dMRI model that employs a multi-compartment framework to indirectly capture aspects of non-Gaussian water behavior but estimates two parameters describing neurite density and orientation dispersion. Though simpler than SMI, the robustness of its microstructural estimates have still been questioned [49], leading to the development of modified models such as C-NODDI, a variant of the original NODDI framework that introduces additional model constraints. This approach has been shown to have improved estimation stability and enhanced sensitivity across various brain regions and conditions [50], and indeed outperformed its respective NODDI-derived parameters in this study. Nonetheless, its individual performance was relatively low compared to MAP, likely as a potential limitation of NODDI’s lower specificity to differences in myelin content, which could diminish its ability to detect and classify nuanced myelin degradation in MCI.

CSF biomarkers have also long been considered important indicators of AD pathology, and our results confirm their relevance in MCI classification. Among the biomarkers analyzed, *pTau/Aβ42* emerged as the only biomarker with statistically significant group differences and had the most discriminative AUC and performance results, suggesting its importance as a marker of dementia pathophysiology. Interestingly, other CSF biomarkers—including *Aβ42*, *Aβ40*, *Tau*, and *pTau*—did not show statistically significant differences between CN and MCI despite their fair AUC performance. This discrepancy could be due to our relatively low CSF sample size or variability in these biomarker levels at the specific MCI stage. It is important to note that the best-performing CSF biomarker, *pTau*/*Aβ42*, achieved an AUC slightly lower than the top-performing dMRI parameter, MAP-*RTAP*, but comparable sensitivity and specificity. Given that CSF extraction is an invasive procedure that some patients may prefer to avoid, these results propose non-invasive dMRI as a viable alternative that provides both comparable diagnostic accuracy and the added benefit of brain region-specific spatial information.

### ***4.2.*** Biomarker Integration

A key finding of this study is that combining dMRI and CSF biomarkers significantly enhanced classification performance as compared to single-modality models. This improved performance suggests that dMRI and CSF offer complementary insights into MCI pathology, potentially due to dMRI reflecting microstructural integrity and neurodegeneration while CSF biomarkers indicate protein dysregulation. Specifically, the top-performing combined model, C-NODDI-*C-NDI + Aβ42/Aβ40*, offered a well-balanced performance of high sensitivity and specificity, providing a potentially useful standalone clinical diagnostic tool. Additionally, since other dMRI and CSF biomarker combinations optimize either sensitivity or specificity—such as C-NODDI-*C-NDI* + *Tau* achieving 93% sensitivity and MAP-*RTAP* + *Tau* achieving 88% specificity—using multiple dMRI classifiers together in a clinical setting could enhance diagnostic accuracy and minimize misclassification. Importantly, while MAP-derived parameters dominated the top individual classifiers, the combined dMRI + CSF classifiers incorporating C-NODDI*-C-NDI* now exhibited the strongest performance across all metrics while exhibiting higher accuracy as compared to previous works [51]. This could potentially be explained by the fact that MAP-derived parameters have been shown to more closely correlated with CSF biomarkers than NODDI-derived parameters [29], and thus, NODDI-derived parameters might benefit more from the addition of CSF information. When used in synchrony, NODDI parameters provide exclusive information on the physical integrity of brain tissue, while CSF biomarkers reflect the biochemical changes associated with Alzheimer’s pathology. This complementary pairing allows for a comprehensive multimodal assessment of AD pathology, including neuronal injury and degeneration, amyloid-β accumulation and tau protein phosphorylation, neuroinflammation, and oxidative stress, resulting in greater classification performance. Further investigation into these synergistic effects may shed new insights into the relationship between microstructural, composition, and biochemical markers of neurodegeneration, and reveal differential clinical applications of the various dMRI models explored in this study.

### ***4.3.*** Machine Learning Interpretability

Model explainability analyses highlight the significance of regional dMRI features and provide insight into the factors driving classification predictions. In the best-performing combined models, ROI values from the frontal, temporal, and parietal lobes emerged as some of the most influential predictors, with lower feature values associated with a greater likelihood of MCI classification. This pattern aligns with the biochemical and theoretical underpinnings of dMRI parameters, reinforcing the model’s sensitivity and validity. For example, lower C-NODDI-*C-NDI* values, which reflect lower axonal density or integrity, are associated with MCI in our models, supporting research evidence that lower neuron density is linked to neurodegeneration in AD patients [52], while MAP-*RTAP*, a measure of water diffusion restriction along the radial direction, supports work indicating microstructural degradation within white matter pathways of MCI participants [53]. These findings additionally support established neuropathological evidence that the frontal, parietal, and temporal lobes are generally impacted in early neurodegeneration [54, 55].

CSF biomarkers, while less influential than dMRI measures in this study, contributed additional predictive value consistent with established findings. A lower *Aβ42*/*Aβ40* ratio was associated with a higher likelihood of MCI classification, reflecting the well-documented role of amyloid pathology in early AD [56]. In contrast, elevated *pTau*/*Aβ42* and *Tau* levels were linked to MCI, reinforcing their significance as indicators of neurodegeneration and aligning with their established association with neurofibrillary tangle accumulation and neuronal injury [57, 58].

The demographic variables generally had a moderate impact on classification, with sex, age, and YoE roughly ranked in order of importance. Male sex was predominantly associated with MCI, agreeing with prior studies indicating a higher MCI risk in males [59, 60], and supported by the statistically significant sex distribution difference in our dataset (*p* = 0.011). The relationship between age and classification was less straightforward—both younger and older individuals were classified as CN, while a subset of younger individuals had a higher likelihood of being classified as MCI. This pattern diverges from the well-established link between increasing age and MCI/AD risk [61]. However, given that age was only close to significance (p=0.1) between the two groups and the nonlinearity of XGBoost decision trees, these findings may reflect hidden confounding or interaction effects, where age influences classification in conjunction with factors such as biomarker levels and cognitive reserve. YoE, which was seen to be nearly significant (*p* = 0.055) between the two groups, played a straightforward but minimized role in classification. Higher YoE was generally associated with CN classification, aligning with the cognitive reserve hypothesis— which proposes that greater educational attainment enhances neural efficiency and offers resilience against neurodegeneration [62]. However, its overall effect on model predictions was still far less significant than that of dMRI and CSF biomarkers. Therefore, educational years likely provide some protective correlation, but structural and molecular markers remain as far stronger predictors of MCI classification.

### ***4.4.*** Limitations and Future Directions

Our study comes with limitations. First, the moderate sample size of the diffusion-weighted ADNI cohort may affect the generalizability of our findings. While model performance was evaluated using 5-fold cross-validation, a larger dataset would allow for a more robust train/validation/test split. Additionally, the ADNI cohort is predominantly composed of non-Hispanic white participants, leading to an imbalanced racial distribution. As a result, race was omitted from classification analysis, and future studies with more diverse populations should explore the potential classification benefits of incorporating this demographic variable. Another limitation of our study is the disparity in sample sizes between dMRI and CSF biomarker data. While our results highlight the diagnostic value of combining dMRI and CSF biomarkers, direct comparisons between single-modality and multimodal models are slightly confounded by the smaller CSF sample size. Future studies with more balanced data across modalities would allow for a clearer evaluation of their relative impact. Next, our classification models were based on ROI-level dMRI features, which aggregate regional information but do not fully capture the voxel-wise spatial detail present in the original parameter maps. This may limit classification performance, and future work should explore deep learning approaches that leverage the full spatial information of dMRI data for improved predictive accuracy. Finally, this study focused on cross-sectional classification of MCI. By including all individuals with MCI, some of whom may not progress to AD, there is added complexity in interpreting diagnostic usefulness, as the findings may not be solely reflective of AD pathology and may blur the distinction between those with early AD and those with other non-AD causes of cognitive decline. Future research should investigate whether these biomarkers can indeed predict longitudinal progression from MCI to AD, further establishing their clinical utility in disease monitoring and prognosis.

## 5. CONCLUSIONS

This study evaluates the diagnostic performance of different advanced dMRI-based models in WM, and emphasizes the complementary roles of these dMRI and CSF biomarkers in detecting neurodegenerative processes. Additionally, it highlights the utility of explainability in machine learning models and provides valuable insights from imaging and fluid information supporting early AD detection and prognosis.

## Data Availability

All processed data produced in the present study are available upon reasonable request to the authors and the ADNI committee.

https://adni.loni.usc.edu/

## 6. ACKNOWLEDGEMENT

This work was supported by the Intramural Research Program of the National Institute on Aging of the National Institutes of Health.

Data collection and sharing for this project were supported by the Alzheimer’s Disease Neuroimaging Initiative (ADNI) through funding from the National Institutes of Health (Grant U01 AG024904) and the Department of Defense (DOD ADNI, award number W81XWH-12-2-0012). ADNI is primarily funded by the National Institute on Aging and the National Institute of Biomedical Imaging and Bioengineering, with additional support from the Canadian Institutes of Health Research for clinical sites in Canada. Private sector contributions are facilitated by the Foundation for the National Institutes of Health (www.fnih.org).

Generous financial support has also been provided by various organizations, including AbbVie, the Alzheimer’s Association, the Alzheimer’s Drug Discovery Foundation, Araclon Biotech, BioClinica, Inc., Biogen, Bristol-Myers Squibb Company, CereSpir, Inc., Cogstate, Eisai Inc., Elan Pharmaceuticals, Inc., Eli Lilly and Company, EuroImmun, F. Hoffmann-La Roche Ltd. and its affiliate Genentech, Inc., Fujirebio, GE Healthcare, IXICO Ltd., Janssen Alzheimer Immunotherapy Research & Development, LLC, Johnson & Johnson Pharmaceutical Research & Development LLC, Lumosity, Lundbeck, Merck & Co., Inc., Meso Scale Diagnostics, LLC, NeuroRx Research, Neurotrack Technologies, Novartis Pharmaceuticals Corporation, Pfizer Inc., Piramal Imaging, Servier, Takeda Pharmaceutical Company, and Transition Therapeutics.

The Northern California Institute for Research and Education serves as the grantee organization, while the Alzheimer’s Therapeutic Research Institute at the University of Southern California coordinates the study. ADNI data are disseminated by the Laboratory for Neuro Imaging at the University of Southern California.

## 7. SUPPLEMENTARY FIGURES

